# Low Pass Whole Genome sequencing - a cost-effective new technique for the identification of aneuploidies and copy number variations: a study of 1372 clinical samples in an Indian cohort

**DOI:** 10.1101/2025.05.18.25327314

**Authors:** Satish Kariyaiah, Rashmi Rasalkar, Meenakshi Bhat, Avinash Pradhan, Deepika Rani Jena, Smitha C, Manjunath V, A Lenika, Jammula SriGanesh, Monisha Morris, Ravi Gupta, Eswarachari Venkataswamy, Sakthivel Murugan S.M., Ramprasad V.L., Priya Kadam

**Author notes:** Conflict of Interest: No conflict of interest has been declared. Satish Kariyaiah and Rashmi Rasalkar are joint first authors.

## Abstract

**Background:** Identification and confirmation of copy number variation is an important aspect of genetic testing for several prenatal scenarios, such as abnormal maternal serum screening, abnormal ultrasound findings, high risk results on non-invasive prenatal screening as well as postnatal settings such as developmental delay, intellectual disabilities, congenital anomalies and dysmorphism as well as in couples with recurrent miscarriages. Low pass whole genome sequencing (lpWGS) followed by copy number variation (CNV) analysis has shown identification of critical pathogenic/likely pathogenic variants comprehensively with great accuracy and cost-effectiveness. We validated this technique on retrospective routine clinical samples previously assessed by an orthogonal tests, either chromosomal microarray or karyotyping or fluorescence in situ hybridization in a diagnostic laboratory and evaluated its performance.

**Material and Methods:** The validation included 112 clinical samples which showed the following events: 14 aneuploidies, 70 copy number variations of size range (50 kb - 116 Mb), 11 mosaics aneuplodies, 4 mosaics CNVs and 4 triploidies. These validation samples were derived from 21 fetuses (6 chorionic villus biopsies and 15 amniotic fluid samples), 25 abortuses and 66 postnatal samples. The validation was performed at two different resolutions - 1Mb and 50Kb. We then assessed the overall yield of the lpWGS assay on 1,260 clinical samples that comprised 801 fetuses, 346 products of conception/intrauterine fetal demise/stillbirth and 113 postnatal cases.

**Results:** We obtained 100% concordance in detecting full aneuploidies and copy number variations including mosaics and triploidy. The overall success rate of this assay that passed all quality parameters was 99.8%. (1235/1238). The overall diagnostic yield for pathogenic and likely pathogenic variants was 13.6% (168/1235), and specifically 7.3%, 28.8%, 15% for fetuses, products of conception/intrauterine fetal demise/stillbirth and post natal samples respectively. An additional 2.9% (36/1235) of overall samples showed CNVs that were classified as variants of unknown significance.

**Conclusion:** Overall, low pass whole genome sequencing is a sensitive and robust technique for identification of aneuploidies and copy number variations in a clinical setting.

## Introduction

Chromosomal abnormalities in the form of aneuploidies and copy number variations (CNVs) as a likely cause of a specific phenotype are important in both prenatal and postnatal clinical settings Different techniques with increasing resolution and progressive technological advances have been employed over the last few decades in the identification of chromosome aneuploidies and CNVs. These include karyotyping which detectsCNVs with resolution of approximately 5Mb or more aneuploidies, polyploidies, balanced and unbalanced structural rearrangements,, and mosaicism^1^, . Fluorescence in situ hybridization (FISH)^2^, Multiplex ligation probe amplification (MLPA)^3^, quantitative polymerase chain reaction^4^ are targeted approaches for specific abnormalities. Chromosome arrays such as comparative genomic hybridization and single nucleotide polymorphism based array^5^ have enabled analysis of a wider spectrum of abnormalities and with increased resolution. Chromosomal microarray had been established as the gold standard for identification of CNVs for structurally abnormal foetuses, stillbirths, products of conception and for children with intellectual disability and developmental delay by professional societies.^6,7^ Overall, they have a proven greater yield of about 5-15% as compared to karyotyping^8^. However, these array techniques are chip- based and probe dependent, are expensive and have less comprehensive coverage. Emerging studies show that next generation sequencing with low coverage are accurate at CNV detection, low cost and with comprehensive coverage.^9,10^

Low-pass whole genome sequencing (lpWGS) as a method to identify aneuploidies and CNVs is relatively new but the utility has been clearly demonstrated in both prenatal and postnatal scenarios.^11,12,13^ LpWGS refers to shallow whole genome sequencing (WGS) at a lower depth, typically around 0.5X to 5X coverage, as compared to the standard depth of 30X to 50X in traditional WGS. This reduction in coverage significantly decreases the cost and time required for analysis while still providing valuable genomic information. Although CMA is currently the first-line investigation for aneuploidies and CNV identificaton, the lpWGS technique developed on different NGS platforms along with sensitive CNV calling algorithms provides an alternative method to detect aneuploidies and CNVs accurately^11^. An increased yield with lpWGS has been demonstrated in a direct comparison between lpWGS and chromosomal microarray in some studies.^14^ Some of the advantages include lower initial input DNA compared to CMA protocols, non-probe dependency, wider coverage of genome, use of existing NGS equipment/platforms, lower costs and manpower requirements for both wet lab and data-analysis procedures, and high throughput with a shortened turnaround time.^12,13^ A Chinese joint expert group in 2019 recommended the use of lpWGS as first tier diagnostic testing in pregnant women for prenatal diagnosis, especially couples with balanced chromosomal translocations, and/or with pregnancy loss.^15^ Consequently, this technique is being increasingly used in clinical practice in tertiary level Chinese hospitals.^16^ Recently updated guidelines by ACMG on the use of NGS techniques for the identification of structural variants state that lpWGS provides an affordable CNV detection method with a similar diagnostic yield and probable enhanced resolution than microarrays.^17^

This study was aimed at validating the lpWGS technique against a standard tests and evaluating its performance for use in detection of aneuploidies and CNVs. After the validation phase, the study aimed at evaluating the diagnostic yield in 1260 clinical samples.

## Materials and Methods

### Validation cohort

This validation was performed in two phases to assess the ability of lpWGS to identifying aneuploidies and CNVs. In the first phase CNVs greater than 1Mb were validated and in the second phase we assessed the sensitivity of lpWG in detecting CNVs greater than 50Kb and in triploidy identification. Samples with chromosomal mosiacism were included in both phases.

Phase I (0.5X - 0.7X): We used 85 consented retrospective clinical samples with known diagnostic results that included abberations ranging in size from 1.34 to 133 Mb. These included 15 prenatal samples (4 chorionic villus biopsies, 11 amniotic fluid), 17 products of conception and 53 postnatal samples (Figure 1). Of the total, 53 samples were previously investigated by chromosomal microarray (CMA) while 11 were investigated by karyotyping (KT), 15 by karyotyping and FISH for microdeletion syndromes, and 6 others were investigated only by FISH for specific microdeletion syndromes, particularly 22q11.2 microdeletion (Table 1). We created a panel of normal reference using 68 clinical samples with no known clinically relevant CNVs as seen on Affymetrix CytoScan™ Optima array. We evaluated these 10 mosaic samples (Table 1, mosaics), of which consisted of 7 were aneuploidy events and four CNVs with mosaic percentages ranging from 15 - 40% in phase 1 (Figure 1).

**Figure 1:**
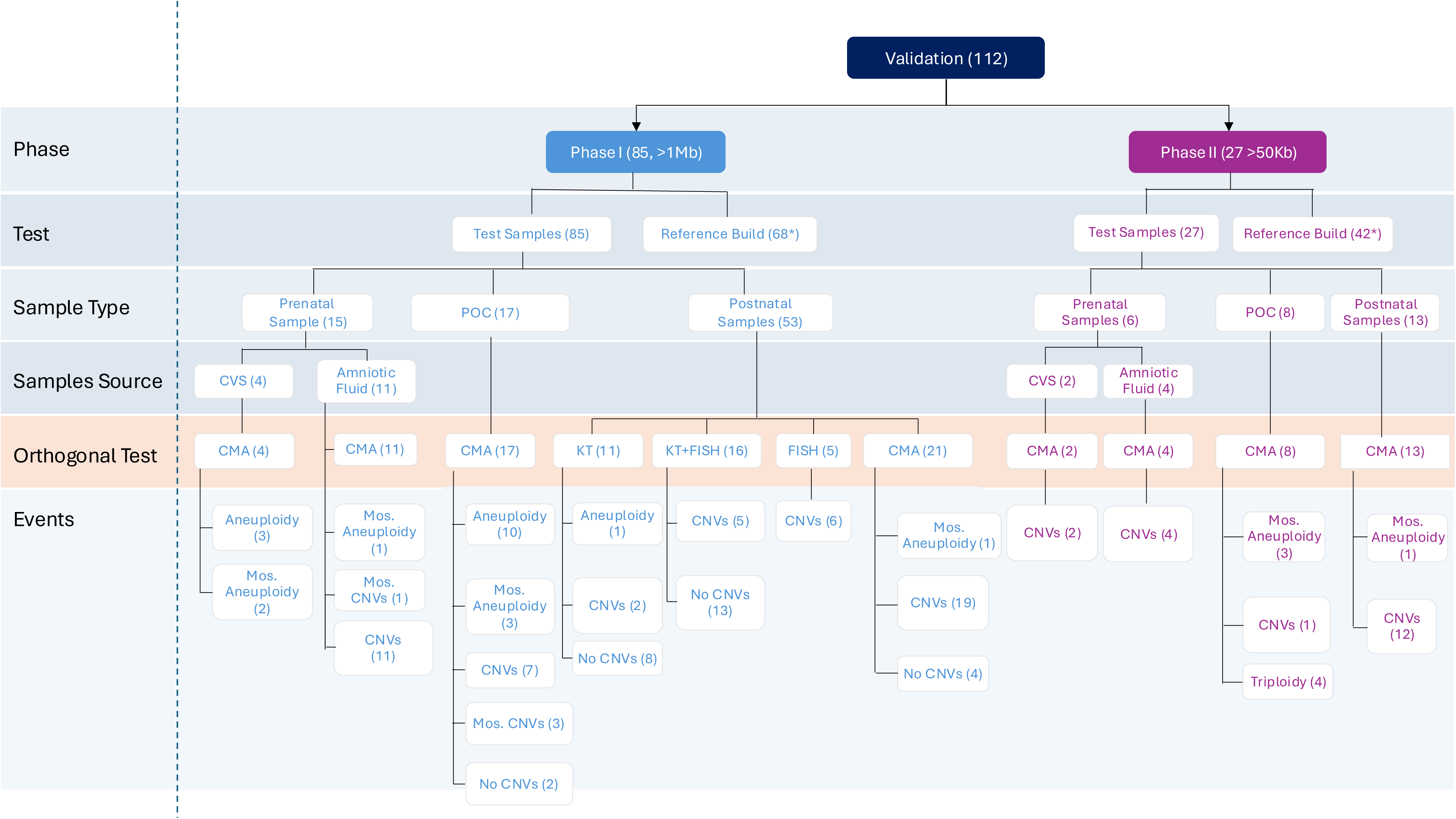
Validation flow chart divided in Phase 1 and Phase II

**Table 1:** List of retrospective clinical samples and its detail used for validation of lpWGS and comparison with Orthogonal test.

| SI No. | Sample Type | Age Range \| Gender | Orthogonal Test | Cytoband with alteration | Orthogonal Test |  |  | Lowpass Whole genome |  |  | Concordance check |
| --- | --- | --- | --- | --- | --- | --- | --- | --- | --- | --- | --- |
|  |  |  |  |  | Start | End | Size (Mb) | Start | End | Size (Mb) |  |
| Phase I |  |  |  |  |  |  |  |  |  |  |  |
| V1 | POC | 1<sup>st</sup> Tri | CMA | Trisomy 21 | 13634137 | 46677460 | 33.04 | 0 | 48129895 | 48.13 | C |
| V2 | CVS | -\|NK | CMA | Trisomy 18 | 136227 | 80255845 | 80.12 | 0 | 78077248 | 78.08 | C |
| V3 | POC | -\|NK | CMA | XXY;<br>Trisomy 13;<br>Trisomy 21 | 251880;<br>18862148;<br>13634137 | 156004066;<br>114342258;<br>46677460 | 155.75;<br>95.5;<br>33.0 | 518401;<br>18875554;<br>13850829 | 155783229;<br>114122514;<br>46484554 | 155.3;<br>95.25;<br>32.63 | C |
| V4 | POC | -\|NK | CMA | XO;<br>Trisomy 18 | 670673;<br>136227 | 156004066;<br>80255845 | 155.33;<br>80.1 | 1;<br>1 | 155270560;<br>80140651 | 155.2;<br>80.14 | C<sup>+</sup> |
| V5 | POC | -\|NK | CMA | XO;<br>Del:2q37.3;<br>Dup:16p13.3p11.2 | 251880;<br>238580161;<br>35881 | 156004066;<br>241841231;<br>32836614 | 155.75;<br>3.26;<br>32.80 | 1<br>238530917;<br>1 | 155270560;<br>242032733;<br>31858519 | 155.2;<br>3.50;<br>31.86 | C<sup>+</sup> |
| V6 | POC | -\|NK | CMA | XXY | 2785592 | 155691858 | 152.91 | 1 | 155270560 | 155.2 | C<sup>+</sup> |
| V7 | POC | -\|NK | CMA | XO | 251880 | 156004066 | 155.75 | 1 | 155270560 | 155.3 | C<sup>+</sup> |
| V8 | POC | -\|NK | CMA | XXX | 2785592 | 155691858 | 152.91 | 518401 | 155783229 | 155.26 | C |
| V9 | POC | 1<sup>st</sup> Tri\|NK | CMA | Trisomy 12 | 64621 | 133201316 | 133.14 | 0 | 133851895 | 133.85 | C |
| V10 | DD | 6-10Y\|F* | KT;<br>CMA | 47,X,der(Y),der(Y)[3<br>2]/46,X,der(Y)[26]/4<br>5,X[2];<br>Dup Yp11.2q11.23 | 2782100 | 26277778 | 23.5 | 1 | 59373566 | 59.3 | C |
| V11 | PB | 6-10 Y \|M | CMA | Dup:Xq28 | 153063080 | 154164940 | 1.1 | 153029585 | 154403555 | 1.37 | C |
| V12 | AF | 2nd Tri\|NK | CMA | Del:Xp22.31 | 6568449 | 8188133 | 1.62 | 6567032 | 8126043 | 1.56 | C |
| V13 | PB | 6-10 Y \|M | CMA | Del:1p36.33p36.32 | 914087 | 4687603 | 3.77 | 1035549 | 4106654 | 3.07 | C |
| V14 | AF | 2nd Tri\|NK | CMA | Del:3q29 | 193054506 | 195291140 | 2.24 | 192701721 | 195307011 | 2.61 | C |
| V15 | DD | 0-5 Y \|M | F-22q;<br>CMA | Negative;<br>Del 3p26.3p26.1;<br>Dup 9q32q34.3 | 20214;<br>114420869 | 4426396;<br>138125937 | 4.4;<br>23.7 | 0;<br>117178796 | 4560000;<br>141213431 | 4.5,<br>24 | C |
| V16 | PB | 0-5 Y \|M | CMA | Dup:3q13.2q13.31 | 112451525 | 115722398 | 3.27 | 112416860 | 115710247 | 3.29 | C |
| V17 | DD | 0-5 Y \|M | KT;<br>F-22q,<br>F-7q;<br>CMA | 46,XY, der(6);<br>Negative;<br>Negative;<br>Dup 3q25.32q29 | 157312388 | 193491389 | 36.2 | 157001502 | 193501721 | 36.5 | C |
| V18 | DD | 0-5 Y \|M | KT;<br>CMA | 46, XY? der (15);<br>Dup 4p16.3p15.2 | 68454 | 25612567 | 25.5 | 0 | 25640993 | 25.4 | C |
| V19 | PB | 0-5 Y \|F | CMA | Del:4q34.1q35.2 | 172079686 | 190036305 | 17.96 | 172151582 | 189871569 | 17.72 | C |
| V20 | DD | 0-5 Y \|NA | F-22q;<br>CMA | Negative;<br>Dup: 4q35.2 | 186249068 | 188459067 | 2.2 | 186981307 | 189181307 | 2.2 | C |
| V21 | PB | 0-5 Y \|F | CMA | Dup:4q25;<br>Del:17p11.2 | 111131505;<br>16880332 | 112863326;<br>20488761 | 1.73;<br>3.61 | 111140415;<br>16916072 | 112967231;<br>20229100 | 1.83;<br>3.31 | C |
| V22 | PB | 0-5 Y \|M | CMA | Del:5q35.2q35.3 | 175989093 | 178055505 | 2.07 | 176170924 | 177632806 | 1.46 | C |
| V23 | AF | -\|NK | CMA | Del:5p15.33p15.2;<br>Dup: 20p13p12.1 | 113462;<br>80929 | 13788173;<br>15514914 | 13.67; 15.43 | 0;<br>0 | 13810000;<br>15360000 | 13.81;<br>15.36 | C |
| V24 | DD | 0-5 Y \|M | KT;<br>F-22q;<br>CMA | 46, XY, der<br>(5)t(5;?)(p15;?);<br>Negative;<br>Del 5p15.33p14.3;<br>Dup 8q23.2q24.3;<br>Dup 6q16.3 | 113462;<br>110872286;<br>100875815 | 23297521;<br>145070385;<br>102907265 | 23.2;<br>34.2;<br>2 | 0;<br>111900277;<br>101330543 | 23313813;<br>146364022;<br>10333054 | 23.3;<br>34.3;<br>2 | C |
| V25 | AF | -\|NK | CMA | Del:6q26q27;<br>Dup:7q33q36.3 | 162350656;<br>133640234 | 170610394;<br>159327017 | 8.26;<br>25.68 | 162025576;<br>133113445 | 171115067;<br>159138663 | 9.09;<br>26.03 | C |
| V26 | PB | 0-5 Y \|M | CMA | Del:7q35q36.3 | 145083337 | 159327017 | 14.24 | 145209927 | 159193037 | 13.98 | C |
| V27 | AF | -\|NK | CMA | Del:7p21.3p15.2 | 10611592 | 26483627 | 15.87 | 10660000 | 26460000 | 15.80 | C |
| V28 | PB | 6-10 Y \|M | CMA | Dup:8q24.21q24.3;<br>Del:21q22.3 | 130350392;<br>46045960 | 145070385;<br>46673449 | 14.72;<br>0.63 | 130269822 | 143035595 | 12.77 | C |
| V29 | AF | -\|NK | CMA | Del:9q22.1q22.31 | 87988200 | 92870072 | 4.88 | 90521336 | 95971336 | 5.45 | C |
| V30 | AF | 2<sup>nd</sup> Tri\|NK | CMA | Del:10p15.3p14 | 100048 | 7701527 | 7.60 | 100048 | 7701527 | 7.60 | C |
| V31 | DD | 0-5 Y \|F | KT;<br>F-22q;<br>CMA | 46, XY, del(10)(p13);<br>Negative;<br>Del 10p15.3p14 | 54087 | 11273168 | 11.2 | 0 | 11460000 | 11.4 | C |
| V32 | POC | -\|NK | CMA | Dup:11p15.5q23.3;<br>Del:22q11.1q11.21 | 230616;<br>16408174 | 116744147;<br>20291006 | 116.51; 3.88 | 230616;<br>16408174 | 116744147;<br>20291006 | 116.51;<br>3.88 | C |
| V33 | PB | 0-5 Y \|M | CMA | Del:13q22.2q31.3 | 75664252 | 91427209 | 15.76 | 75526376 | 91390174 | 15.86 | C |
| V34 | POC | -\|NK | CMA | Del:13q32.3q34;<br>Dup:16p13.3p13.2 | 101067948;<br>35881 | 114342258;<br>8831829 | 13.27;<br>8.79 | 101770197;<br>5260000 | 115169878;<br>8910000 | 13.40;<br>3.65 | C |
| V35 | AF | 2<sup>nd</sup> Tri\|NK | CMA | Del:15q13.2q13.3 | 30820717 | 32622039 | 1.80 | 30885000 | 32485000 | 1.60 | C |
| V36 | PB | 0-5 Y \|M | CMA | Del:15q11.2q13.1 | 22582283 | 28697883 | 6.12 | 23571052 | 28127176 | 4.56 | C |
| V37 | DD | 6-10 Y \|M | ANGL :<br>CMA | Negative;<br>Del 17p12 | 14196248 | 15601354 | 1.4 | 14000000 | 15500000 | 1.4 | C |
| V38 | AF | 2<sup>nd</sup> Tri\|NK | CMA | Del:17q24.1q24.2 | 66060439 | 67467739 | 1.41 | 66032271 | 67400943 | 1.37 | C |
| V39 | DD | 0-5 Y \|F | KT;<br>CMA | 46,XX;<br>Dup 17q11.2 | 30689342 | 31870304 | 1.2 | 29 007057 | 30707057 | 1.7 | C |
| V40 | PB | 0-5 Y \|M | CMA | Del:18q21.2q21.33 | 54720552 | 62003701 | 7.28 | 54761202 | 61988318 | 7.23 | C |
| V41 | POC | -\|NK | CMA | Dup:19q13.2 | 39151229 | 40281520 | 1.13 | 38942109 | 40426354 | 1.48 | C |
| V42 | PB | 50-55 Y \|F | CMA | Del:21q22.3 | 45329955 | 46673449 | 1.34 | 45373861 | 46598733 | 1.22 | C |
| V43 | PB | 6-10 Y \|M | CMA | Del:21q11.2q21.3 | 13644166 | 25925065 | 12.28 | 13850829 | 25933790 | 12.08 | C |
| V44 | DD | 26-30 Y \|F | F-22q | Positive | - | - | - | 18862659 | 23270380 | 4.4 | C |
| V45 | DD | 0-5 Y \|F | F-22q;<br>CMA | Positive;<br>Del 22q11.21 | 18950000 | 21108369 | 2.2 | 18862659 | 21470380 | 2.6 | C |
| V46 | PB | 11-15 Y \|M | CMA | Del:22q13.32q13.33 | 48199706 | 50759338 | 2.5 | 48199706 | 50759338 | 2.6 | C |
| V47 | PB | 0-5 Y \|M | CMA | Del:3p25.3;<br>Dup:15q11.2 | 8617702;<br>22582283 | 9971426;<br>23060000 | 1.35;<br>0.48 | 8392526;<br>22852677 | 10442526;<br>23095635 | 2.05;<br>0.24 | C |
| V48 | PB | 0-5 Y \|F | CMA | Del:10q22.3;<br>Del:15q11.2q13.1 | 79625681;<br>23102621 | 80213053;<br>28458904 | 0.59;<br>5.36 | 79551950;<br>23571052 | 80232513;<br>28127176 | 0.68;<br>4.56 | C |
| V49 | POC | -\|NK | CMA | Mos. Trisomy<br>13[0.35] | 18862148 | 114342258 | 95.5 | 18800001 | 114400000 | 95.6 | 2GB |
| V50 | PB | 0-5 Y\|NK | CMA | Mos. Monosomy X[0.3] | 2785592 | 155691858 | 152.91 | 200001 | 156000000 | 155.8 | 2GB |
| V51 | POC | -\|NK | CMA | Mos. Dup:Xp22.31q21.1[0.4];<br>Mos.Del:Xq21.1q28[0.4] | 8186478;<br>78579045 | 78576179;<br>155233098 | 70.39;<br>76.65 | 0;<br>76708058 | 9150000;<br>155270560 | 42.2;<br>78.6 | 2GB |
| V52 | POC | -\|NK | CMA | Mos. Monosomy 19 [0.3] | 260912 | 58445449 | 58.1 | 200001 | 58600000 | 58.4 | 2GB |
| V53 | POC | -\|NK | CMA | Mos. Trisomy 8 [0.20] | 208049 | 145070385 | 144.8 | 200001 | 145000000 | 144.8 | 2GB |
| V54 | CVS | -\|NK | CMA | Mos. Trisomy 13 [0.15] | 18862148 | 114342258 | 95.5 | 18800001 | 114400000 | 95.6 | 2GB |
| V55 | AF | 2<sup>nd</sup> Tri\|NK | CMA | Mos. Trisomy 22[0.20] | 16408174 | 50759338 | 34.3 | 15800001 | 50800000 | 35.0 | 2GB |
| V56 | CVS | 2<sup>nd</sup> Tri \|NK | CMA | Mos. Trisomy 20 [0.30];<br>Trisomy 21 | 81022;<br>13634137 | 64282292;<br>46677460 | 64.2;<br>33.04 | 1;<br>13000001 | 64200000;<br>46800000 | 64.2;<br>33.8 | 2GB |
| V57 | AF | 2<sup>nd</sup> Tri \|NK | CMA | Mos. Dup: 1q21.1q25.2[0.36] | 143459799 | 178083001 | 34.6 | 143200001 | 178200000 | 35.0 | 2GB |
| V58 | POC | -\|NK | CMA | Mos. Del: 7q11.21q21.11[0.4] | 62437390 | 77764079 | 15.3 | 10660000 | 26560000 | 15.9 | 2GB |
| Phase II |  |  |  |  |  |  |  |  |  |  |  |
| V59 | PB | 41-45 Y \|M | CMA | Dup:Xp11.22 | 53522223 | 53604372 | 0.08 | 53531129 | 53596588 | 0.07 | C |
| V60 | PB | 11-15 Y \|M | CMA | Hom.Del:Xp11.23p11.22 | 50034443 | 50205615 | 0.17 | 50046551 | 50203420 | 0.16 | C |
| V61 | AF | -\|NK | CMA | Hom.Del:Xp21.1 | 31613916 | 31761327 | 0.15 | 31618133 | 31746217 | 0.13 | C |
| V62 | PB | 0-5 Y \|M | CMA | Del: 6q25.3 | 156987387 | 157101227 | 0.113 | 157010931 | 157094882 | 0.1 | C |
| V63 | PB | 0-5 Y \|M | CMA | Del:2p16.3 | 47783201 | 47837567 | 0.05 | 47777481 | 47847637 | 0.07 | C |
| V64 | CVS | -\|NK | CMA | Del:2q31.2 | 178716155 | 178979102 | 0.26 | 178697107 | 178972060 | 0.27 | C |
| V65 | POC | -\|NK | CMA | Del:16p11.2 | 29555976 | 30215609 | 0.66 | 29600001 | 30200000 | 0.59 | C |
| V66 | PB | 0-5 Y \|F | CMA | Del:6p25.3 | 1593382 | 1701415 | 0.11 | 1585474 | 1692283 | 0.11 | C |
| V67 | PB | 0-5 Y \|M | CMA | Del:9q34.3 | 137547852 | 137657680 | 0.11 | 137525141 | 137651995 | 0.13 | C |
| V68 | CVS | -\|NK | CMA | Del:10p13 | 14944633 | 15022198 | 0.08 | 14945159 | 15033707 | 0.09 | C |
| V69 | PB | 16-20 Y \|F | CMA | Del:14q23.1 | 58280647 | 58458765 | 0.18 | 58278240 | 58462794 | 0.18 | C |
| V70 | DD | 6-10 Y \|M | CMA | Del:14q22.1 | 50553477 | 50726580 | 0.17 | 50561349 | 50720465 | 0.16 | C |
| V71 | AF | -\|NK | CMA | Del:15q11.2 | 22582283 | 23102621 | 0.52 | 22781078 | 23102105 | 0.32 | C |
| V72 | PB | 0-5 Y \|M | CMA | Del:15q25.2 | 82536429 | 82583477 | 0.05 | 82547192 | 82615826 | 0.07 | C |
| V73 | DD | -\|NK | CMA | Del: 6q27 | 170279913 | 170605209 | 0.32 | 170267801 | 170601115 | 0.3 | C |
| V74 | PB | 0-5 Y \|M | CMA | Del:16p13.3 | 2081121 | 2235560 | 0.15 | 2092639 | 2228021 | 0.14 | C |
| V75 | PB | 0-5 Y \|M | CMA | Dup:17q21.31 | 46110932 | 46215310 | 0.10 | 46087375 | 46286901 | 0.20 | C |
| V76 | AF | -\|NK | CMA | Del:22q11.21 | 20704600 | 21110475 | 0.41 | 20725934 | 21092778 | 0.37 | C |
| V77 | PB | 6-10 Y \|F | CMA | Del:22q13.33 | 50707646 | 50818468 | 0.11 | 50710333 | 50730443 | 0.11 | C |
| V78 | POC | -\|NK | CMA | Mos.Trisomy 16 [0.55] | 35881 | 90088654 | 90.05 | 40001 | 90120000 | 90.1 | C |
| V79 | POC | -\|NK | CMA | Mos.Trisomy 13<br>[0.62] | 18862148 | 114342258 | 95.48 | 18875194 | 114345127 | 95.5 | C |
| V80 | PB | 0-5 Y \|F | CMA | Mos.Trisomy 9 [0.5] | 203862 | 138125937 | 137.92 | 100001 | 138160000 | 138.1 | C |
| V81 | POC | -\|NK | CMA | Mos.Monosomy<br>21[0.3] | 13634137 | 46677460 | 33.04 | 12940001 | 46680000 | 33.7 | C |
| V82 | POC | -\|NK | MCC | Triploidy | - | - | - | - | - | - | C |
| V83 | POC | -\|NK | MCC | Triploidy | - | - | - | - | - | - | C |
| V84 | POC | -\|NK | MCC | Triploidy | - | - | - | - | - | - | C |
| V85 | POC | -\|NK | QF-PCR | Triploidy | - | - | - | - | - | - | C |

CVS – Chorionic Villous Biopsy, PB – Peripheral Blood, POC – Products of Conception, DD – Direct DNA, AF – Amniotic Fluid, 1<sup>st</sup> Tri – First Trimester, 2<sup>nd</sup> Tri – Second Trimester, NK- Not Known, CMA – Chromosomal Microarray, MCC – Maternal Cell contamination, QF-PCR – Quantitative Fluorescence – Polymerase Chain Reaction
**Note:** Control samples were used to build the reference model, Phase I – 68 normal samples and Phase II – 42 normal samples, other samples where no event was detected by orthogonal testing - 6 CMA, 8 karyotyping, 13 karyotyping and FISH, samples are not shown in the Table 1, C - Concordance between CMA data and IpWGS by either Wiseconder/DRAGEN CNV calling tools.
<sup>†</sup>DRAGEN is capable of detecting events such as segmental loss or gain through its CNV calling workflow. However, it does not directly provide information on sex chromosome anomalies. To address this, we utilized our internal workflow to compute z-scores for the X chromosome by comparing the sample against control samples. Based on these z-scores, we predict whether the sample shows signs of conditions such as XO or XXY. Samples V47 and V48 in phase I had variants below the predetermined threshold.

Phase II (3-5X Depth): To increase the resolution for identifying smaller CNVs (up to ∼50Kb) as well as mosaicism and triploidy, which require greater sequencing depth to capture smaller variants, a total of 27 consented retrospective clincial samples were included (6 prenatal samples, 8 products of conception, and 13 postnatal samples). All these samples were previously investigated by either Affymetrix CytoScan™ Optima array, CytoScan™ 750K Array or CMA CytoScan™ XON Array and had CNVs of size range less than 1Mb and greater than 50 Kb (range 47 - 660 Kb) (Table 1, Phase II, Figure 1). To build a reference set, 42 samples without CNVs on CytoScan™ 750K Array were included.

We also included four triploidy samples, identified by MCC testing (16 STR markers/QF-PCR for aneuploidy) and CMA by a inhouse triploidy detection algorithm in phase II. We also included four mosaic aneuploidies with mosaic percentages from 30 - 65% (Table 1).

### Library preparation and sequencing

DNA was extracted using both automated (QiaSymphony) and manual methods. QC-passed samples were used for whole genome library preparation at a minimum of 50ng on the Agilent Bravo platform using commercially available library prep kits. Prepared libraries were sequenced on Illumina platforms (NovaSeq 6000 or X Plus). Around 2Gb data per sample was generated in Phase I (∼ 1 Mb resolution) and 15 Gb data for the samples used for ∼50 Kb resolution in Phase 2. The sequencing results generated ∼16 million paired-end reads of 150 bp per sample, corresponding to a 0.5X - 0.7X for >1Mb size and 3X - 5X coverage for >50 Kb size.

### Bioinformatics analysis and variant classification

The raw paired-end FASTQ files were processed using Cutadapt to remove adapter sequences, reads shorter than 20 bp or with a quality score below 10 phred score. CNV analysis was performed using both WisecondorX and Illumina’s DRAGEN v4.2. WisecondorX was employed solely for validation purposes.

For WisecondorX analysis, the trimmed reads were aligned to the GRCh38 reference genome using the Sentieon BWA aligner (sentieon-genomics-201808). Coverage at a defined bin size was computed from the aligned BAM files for each sample and converted into a WisecondorX compatible NPZ format. All reference NPZ files were subsequently merged to create a final reference NPZ file. Segmentation was then performed based on reference ploidy, grouping bins with similar ploidy levels. Segments showing ploidy deviations from the reference were identified and classified as copy number variations.

For analysis using DRAGEN, trimmed reads were directly employed for alignment and CNV detection. The reads were aligned to the GRCh38 reference genome. CNV calling was performed through a comparative coverage-based approach, where coverage across predefined bin sizes was calculated. Subsequently, the data underwent bias correction, addressing GC-content bias, resulting in the generation of a corrected target counts file. Normalization was then carried out ensuring that all reference files were also GC-corrected if the sample had undergone GC bias correction. Once normalized, segmentation of the data was performed based on the processed signal. This was followed by CNV detection, where any segment displaying a significant deviation from the normal ploidy level—established using the reference—was classified as a CNV event.

To identify larger events, a bin size of 200 Kb was used, with analysis performed against a model based on 68 reference samples. Similarly, for identifying events larger than 50 Kb, a finer bin size of 20 Kb was applied, utilizing a model constructed from 42 reference samples.

Triploidy was identified by evaluating the proportion of mutant reads in heterozygous SNPs.^18^ To perform this analysis, high-quality heterozygous SNPs from the 1,000 Genomes Project were utilized. For each site with sufficient depth, allele frequencies were calculated based on reference and alternate bases across all samples. In triploidy, the allele frequency at heterozygous positions is expected to be around 0.33 or 0.66, indicating a genetic gain. The variants were then categorized into three distinct groups according to their copy number states: 0.33 (with a range of 0.28–0.38), 0.66 (range of 0.62–0.72), and 0.5 (range of 0.45–0.55). Following this, the ratio between the number of sites with 0.33 and 0.66 frequencies, relative to those with 0.5, was computed. Upon comparing these ratios between control and positive samples, a clear distinction emerged. Samples exhibiting a ratio greater than 2.4 were classified as positive for triploidy.

Annotation and classification of all CNVs was performed using an in-house workflow developed based on the ACMG and Clinical Genome Resource (ClinGen) guidelines.

### Diagnostic yield of serial clinical samples by lpWGS in a clinical setting

We evaluated the assay yield from 1,260 clinically consented samples (prenatal - 801, POC/Intrauterine deaths/stillbirths - 346, postnatal - 113). Demographic details such as maternal age, gestational age and clinical indications for testing were recorded. All prenatal samples were subjected to maternal cell contamination (MCC) analysis either directly or combined with aneuploidy testing through quantitative fluorescence polymerase chain reaction (QF-PCR). MCC and QF-PCR analysis also identified presence of triploidy. Briefly, the Devyser Compact v3 QF-PCR kit was used in accordance with the manufacturer’s protocol. All PCR products were genotyped using a SeqStudio Genetic Analyzer (Thermo Fisher Scientific) with GeneMappe software 6 (Thermo Fisher Scientific). Only MCC negative samples were processed further. These samples were evaluated after the first phase of validation and therefore the CNV resolution was limited to 1Mb.

## Results

We validated the data in two phases, by evaluating the performance of lpWGS 112 samples to detect aneuploidies and CNVs against previous orthogonal data from CMA, karyotype and FISH. Then, we analysed the performace in 1260 consecutive clinical samples.

### Validation results

#### Phase I – Identification of large CNV

All aneuploidies and CNVs greater than 1Mb, identified by orthogonal tests were identified by lpWGS (Table 1).

lpWGS concordance with CMA - All 13 aneuploidies and 37 CNVs identified by previous testing of 53 samples by chrosomsomal microarray were also identified by lpWGS (Table 1). The CNV size ranged from 1.22 – 116.51 Mb. There was a high degree of concordance between the event size detected by CMA and lpWGS as seen in the scatter plot (Figure 2, I). The six samples that did not show CNVs on CMA did not show any CNVs on lpWGS either. To evaluate reproducibility, we re-processed 13 samples three times, all of which showed full concordance (Supplementary Table 1).

**Figure 2:**
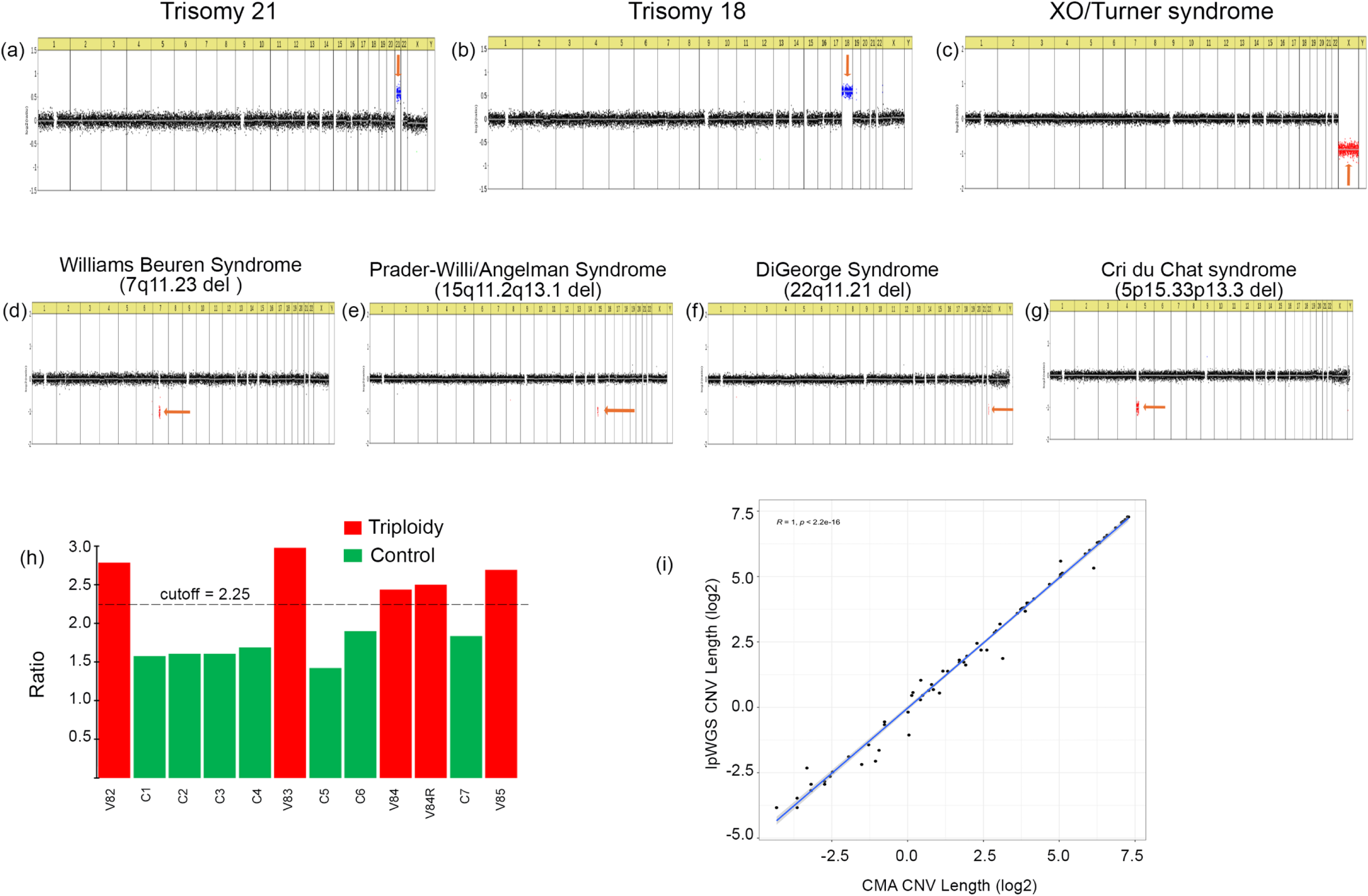
Commonly occurring Aneuploidies and events Trisomy 21(a) Trisomy 18 (b), XO/Turner syndrome (c), 7q11.23 del / Williams Beuren Syndrome (d), 15q11.2q13.1 del / Prader-Willi/Angelman Syndrome (e), 22q11.21 del / DiGeorge Syndrome (f), 5p15.33p13.3 del / Cri du Chat syndrome (g), Triploidy (h), scatterplot CMA vs lpWGS (i)

lpWGS concordance with Karyotyping data – Of eleven samples initially tested by karyotyping, two abnormal events, one each in V10 and V18 had been identified by karyotype. LpWGS identified and further characterized these two events (Table 1), first, V10, a mosaic karyotype involving derivative of Y chromosome was recognized as four copies of Chromosome Y, second, V18, karyotype had shown 46,XY,?der(15), lpWGS revealed a duplication of 25.4 Mb at 4p16.3-p15.2, identifying the probable chromosomal content within the derivative chromosome. Additionally, lpWGS identified a CNV, duplication of size 1.7 Mb at cytoband region 17q11.2, in sample V39, where the karyotype had been reported as normal. However, this was not expected to be detected on karyotype since it was below the detection limit. These three events were further confirmed by chromosomal microarray (Table 1). The remaining eight samples that had not show any abnormality on karyotype with a detection limit of 5Mb, did not show any CNV on lpWGS of size greater than 5Mb, either.

lpWGS concordance with Karyotyping and FISH for 22q11.2 microdeletion data – Of the sixteen samples previously evaluated by both karyotyping and FISH for 22q11.2 microdeletion syndrome, 3 samples, V24, V17 and V31, had shown abnormalities (Table 1). LpWGS assay identified 5 CNVs among these three samples. Three CNVs were seen in one sample alone (V24). This case had a karyotype of 46,XY,der(5)t(5;?)(p15;?). lpWGS identified, a deletion at 5p15.33-p14.2 and duplications at 8q23.2-q24.3 and 6q16.3, with sizes of 23.3 Mb, 34.3 Mb, and 2 Mb, respectively. Parental karyotyping was recommended due to suspected structural variation, however, no further information was available as the case was lost to follow-up. These events were further confirmed by running a CMA (Table1).

Second, sample V17 had shown a karyotype of 46,XY,der(6) and no deletion identified by FISH probe 22q11.2 deletion or for 7q deletion. However, lpWGS identified a 36.5 Mb duplication at cytoband 3q25.32-q29, clarifying the chromosomal component of the derivative chromosome while no variant was seen in q arm of either chromosome 7 or 22. Lastly, in V31, where karyotype had identified an deletion, 46,XY,del(10)p13) and where FISH was negative for 22q11.2, lpWGS identified the precise breakpoints and size, at cytoband region 10p15.3-p14 and size of 11.4Mb respectively. Both findings of lpWGS were again confirmed on CMA (Table 1). In the 13 remaining samples, where no event was seen by karyotyping and FISH, lpWGS also did not identify any CNV.

lpWGS concordance by FISH alone - We evaluated 5 samples by lpWGS that were previously evaluated by FISH, four by FISH for 22q11.2 deletion, and one for 15q11 deletion for Angelman syndrome alone, based on clinical presentation. Two samples that had shown the 22q11.2 microdeletion by FISH were also identified by lpWGS where microdeletions in cytoband region 22q11.21 of sizes 4.4 (V44) and 2.6 Mb (V45) were identified. For a sample investigated by FISH for Angelman syndrome, V37, no variation was detected on chromosome 15; however, lpWGS identified a 1.4 Mb deletion at 17p11.2. The events seen in V37 and V45 was also confirmed by chromosomal microarray (Table 1). However V44 could not be confirmed as the sample was exhausted.

In two samples, that did not show any deletion by FISH for 22q11.2 deletion, showed two CNVs in V15, a terminal gain and loss, at cytoband region 9q32-q34.3 and 3p26.3 - p26. of size 24 and 4.5 Mb respectively and in the other sample, V20, one CNV, a duplication at 4q35.1-q35.2 of size 2.2Mb was detected. The events in V15 and V20 were confirmed on CMA (Table 1)

Concordance of mosaic events – Of the 10 mosaic samples analyzed in Phase I, the seven mosaic aneuploidies that showed mosaicism between 15% - 35% demonstrated on CMA data showed concordance with lpWGS in terms of relative size and coordinates of chromosomal events. Sample V51, had exhibited two distinct events—a gain and a loss on the X chromosome measuring 70.4 Mb and 76.5 Mb, respectively—showed variability in size when analyzed by lpWGS, with the gain measured at 41 Mb and the loss at 9.2 Mb. Of the mosaic CNVs, in three samples that showed about 40% mosaicism, of size range 15.9 –78.6Mb were identified by lpWGS.

Overall, when CMA, karyotype, and FISH results were used as the reference, lpWGS demonstrated, considering at least one CNV calling algorithm, 100% sensitivity for detecting CNVs previously identified, highlighting the high consistency and robust performance of this NGS platform.

#### Phase II – Identification of smaller CNVs, triploidy, mosaics

Of the 27 samples analyzed in Phase 2, all variations identified by CMA were successfully detected by lpWGS. The DRAGEN^TM^ software, which demonstrated the highest concordance with CMA data, was selected for subsequent analysis. The detected CNV size ranged from 70 kb – 880 kb.

Identification of mosaic aneuploidies and CNVs In Phase 2, a different set of mosaic aneuploid samples was analyzed using higher depth. The samples exhibited 30–62% mosaicism, were readily identifiable by both CNV calling tools. The diverse sample set and varied processing conditions allowed for an in-depth assessment of the method’s accuracy and consistency across multiple scenarios.

Identification of Triploidy - All four cases exhibited a significantly higher ratio compared to the controls and were detectable as Triploidy. One sample was reanalyzed for reproducibility, yielding fully concordant results (Figure 2h).

### Clinical Experience with 1260 Samples Demographics and Indications for Testing

The demographic data and diagnostic yield are summarized in Table 2, while clinical indications grouped into broad categories are presented in Figure 3. For the prenatal samples, the most frequent indication for invasive testing was high risk on maternal serum screening (45.4%), followed by ultrasound abnormalities (36.8%). In products of conception (POCs), the leading indications were spontaneous abortion, missed abortion, or intrauterine death (IUD) (42%) followed by ultrasound abnormalities (33.5%). In postnatal cases, dysmorphism and congenital abnormalities being the most common.

**Figure 3:**
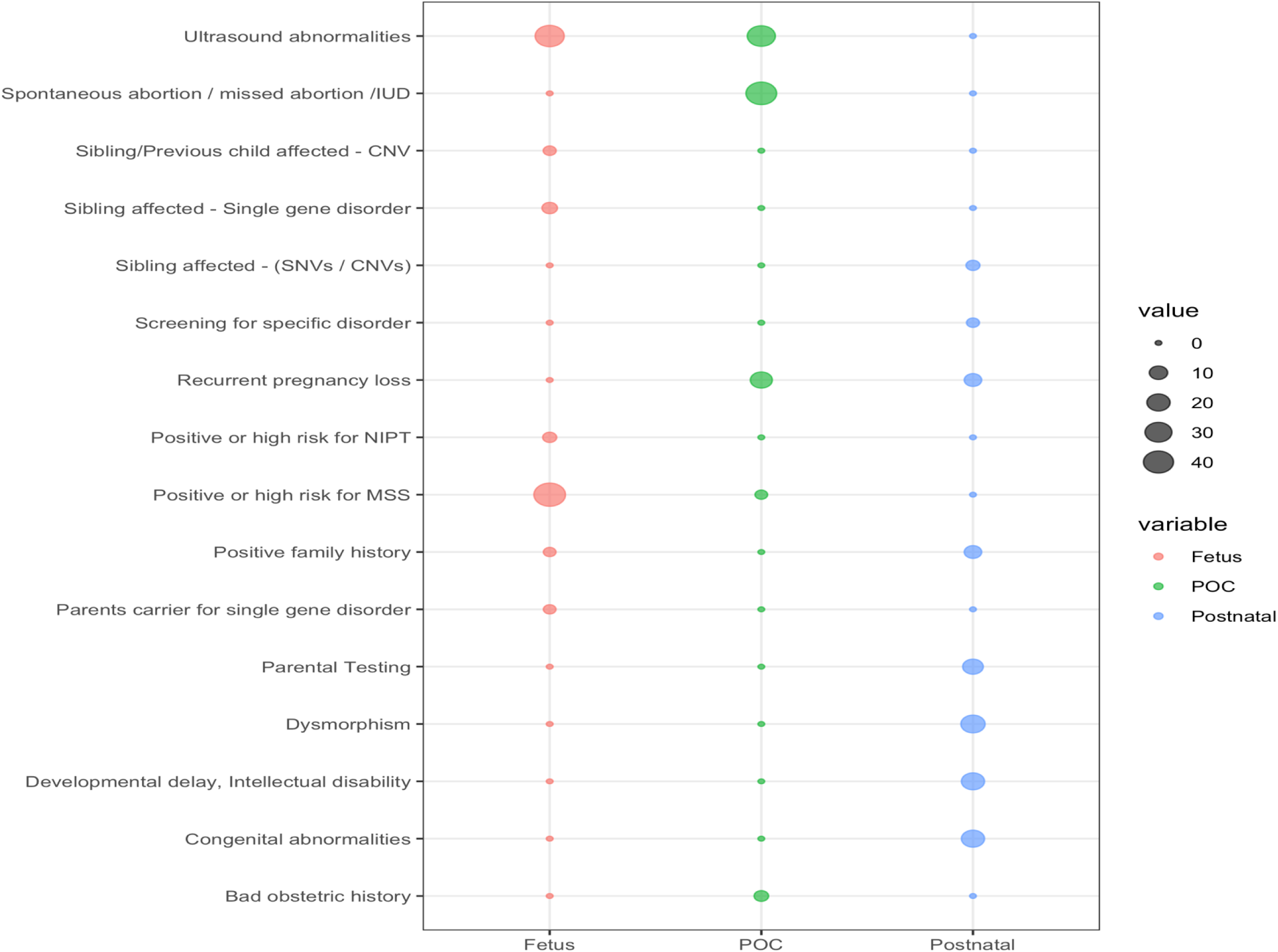
Distribution of Clinical indications of 1260 samples **A**. Fetus, **B**. POC, **C**. Postnatal

**Table 2:** Demographic data and Diagnostic yield of the 1,260 samples evaluated by lpWGS.

| (n=1,260) | Samples | Gestational Age / Age | Maternal Age | Normal | Abnormal |  |
| --- | --- | --- | --- | --- | --- | --- |
|  |  |  |  |  | P/LP | VoUS |
| **Fetus** | 801 (63.6%) | 18W 5D<br>(11W 1D - 33W 3D) | 30.2<br>(17-45) | 90.1% (720/799) | 7.3% (58/799) | 2.6% (21/799) |
| **POC/IUD/Still birth** | 346 (27.5%) | 18W 0D<br>(7W 3D - 34W) | 29.6<br>(18-47) | 68.7% (222/323) | 28.8% (93/323) | 2.5% (8/323) |
| **Postnatal** | 113 (9.0%) | 16Y 2M<br>(10D - 49Y) | - | 78.8% (89/113) | 15.0% (17/113) | 6.2% (7/113) |
| **Overall** | - | - | - | **83.5% (1031/1235)** | **13.6% (168/1235)** | **2.9%(36/1235)** |

### Assay Success Rate and Technical Performance

Result by lpWGS assay was obtained in 99.8% (1235/1238) and therefore failure rate was only 0.2%. Results were obtained in all postnatal samples, with assay failure observed in only three prenatal samples (1 amniotic fluid sample and 2 POCs). The call rate and distribution of normal and abnormal samples by category is shown in Figure 4.

**Figure 4:**
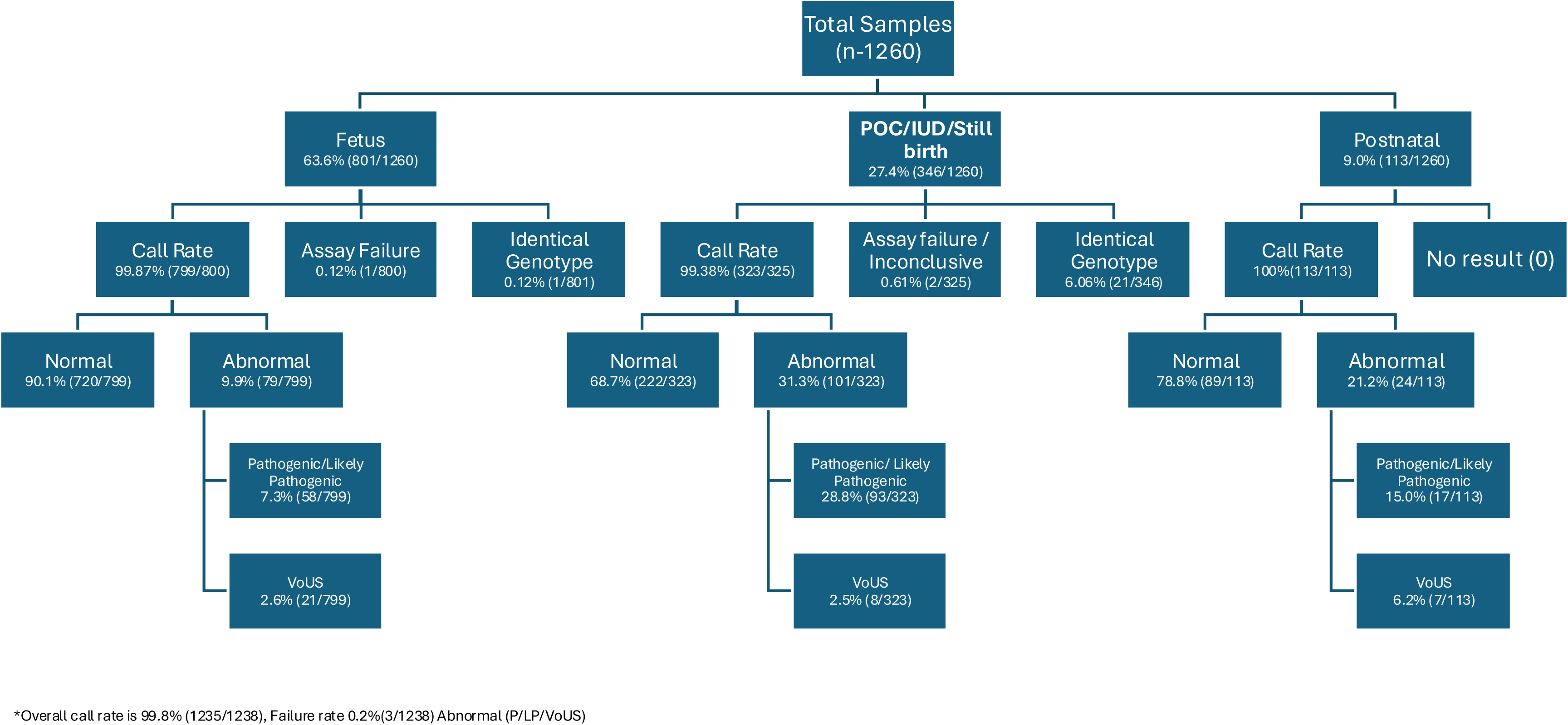
Diagnostic yield for lowpass whole genome sequencing of 1260 samples.

A total of 22 samples (21 POCs and 1 CVS) exhibited identical maternal and fetal genotypes (Figure 4). No fetal genotype was identified by maternal cell contamination (MCC) or quantitative fluorescence PCR (QF-PCR) which might have been caused by improper sample collection. These samples were excluded from further analysis

### lpWGS Concordance with common aneuplodies identified by QF-PCR

Of the 1260 samples, 1147 were prenatal, of which 650 were booked for rapid aneuploidy screening by QF-PCR. Of these, 51/639 (7.9%) aneuploides in chromosomes 21,18,13, and sex chrosomsome aneuplodies were identified by QF-PCR which were also detected by lpWGS. Also in 588/639 (92%) samples where QF-PCR did not pick up any anomaly in these chromosomes showed normal two copies by lpWGS as well, giving 100% concordance for common aneuploidy detection. Additional aneuplodies in chromosomes 3, 14, 16, 22 were identified on lpWGS. The remaning 11 samples showed identical genotype to the mother indicating absence of fetal DNA in the POC tissue and were therefore exluded from further testing by lpWGS.

### Diagnostic Yield

Of the 1,235 samples with identified results (204/1235, 16.5%) 131 were aneuploidies and 73 significant CNVs (Table 3). Of these 37 were pathogenic/likely pathogenic copy number variations (CNVs) and 36 were classified as variants of uncertain significance (VOUS). The overall diagnostic yield for aneuplodies and pathogenic/likely pathogenic CNVs was 13.6%, specifically, x7.3% in fetuses, 28.8% in early miscarriages, intrauterine deaths, and stillbirths, 15% in postnatal cases (Table 2). Variants classified as VOUS accounted for 2.9% of all samples: specifically 2.6% in prenatal cases, 2.5% in POCs/IUDs/stillbirths, 6.2% in postnatal cases (Supplementary Table 2).

**Table 3:** Aneuploidies and pathogenic and likely pathogenic variants identified by LP_WGS. Triploidy was identified by MCC/QF PCR data. Subsequently algorithm was optimized to identify the Triploidy by increasing the depth of sequencing in validation phase II.

| Nomenclature | Overall<br>(n=168) | Fetus<br>(n=58) | POC/IUD/Still birth<br>(n=93) | Postnatal<br>(n=17) |
| --- | --- | --- | --- | --- |
| **Common Aneuploidies** |  |  |  |  |
| Trisomy 21 | 38 (22.6%) | 28 (48.3%) | 8 (8.6%) | 2 (11.8%) |
| Trisomy 18 | 16 (9.5%) | 11 (19.0%) | 5 (5.4%) | - |
| Trisomy 13 | 6 (3.6%) | 3 (5.2%) | 3 (3.2%) | - |
| **Sex Chromosome Aberrations** |  |  |  |  |
| Turner syndrome | 19 (11.3%) | 1 (1.7%) | 18 (19.4%) | - |
| Triple X syndrome | 2 (1.2%) | 2 (3.4%) | - | - |
| Klinefelter syndrome | 2 (1.2%) | 2 (3.4%) | - | - |
| Jacob's syndrome | 2 (1.2%) | 1 (1.7%) | 1 (1.1%) | - |
| Others | 3 (1.8%) | - | 3 (3.2%) | - |
| **Rare Autosomes Aberrations** |  |  |  |  |
| Trisomy 3 | 2 (1.2%) | - | 2 (2.2%) | - |
| Trisomy 7 | 2 (1.2%) | - | 2 (2.2%) | - |
| Trisomy 8 | 2 (1.2%) | - | 2 (2.2%) | - |
| Trisomy 10 | 2 (1.2%) | - | 2 (2.2%) | - |
| Trisomy 14 | 3 (1.8%) | - | 3 (3.2%) | - |
| Trisomy 15 | 2 (1.2%) | - | 2 (2.2%) | - |
| Trisomy 16 | 9 (5.4%) | - | 9 (9.7%) | - |
| Trisomy 17 | 1 (0.6%) | - | 1 (1.1%) | - |
| Trisomy 22 | 7 (4.2%) | - | 7 (7.5%) | - |
| **Other combinations** |  |  |  |  |
| Klinefelter syndrome +Trisomy 18 | 1 (0.6%) | - | 1 (1.1%) | - |
| **Triploidy** | 12 (7.1%) | - | 12 (12.9%) | - |
| **Aneuploidy (Including Triploidy)** | 131 (78.0%) | 48 (82.8%) | 81 (87.1%) | 2 (11.8%) |
| **Copy number variations**<br>**(Pathogenic/Likely pathogenic)** | 37 (22.0%) | 10 (17.2%) | 12 (12.9%) | 15 (88.2%) |
| **<5Mb size** | 20<br>(20/1235,1.6%)<br>(20/168,11.9%) | 7<br>(7/1235,0.6%)<br>(7/58, 12.1%) | 6<br>(6/1235,0.5%)<br>(6/93,6.5%) | 7<br>(7/1235,0.6%)<br>(7/17,41.2%) |
| **>5 Mb size** | 17<br>(17/1235,1.4%) | 3<br>(3/1235,0.2%) | 6<br>(6/1235,0.5%) | 8<br>(8/1235,0.6%) |

Aneuploidies - Aneuploidies accounted for 78% (131/168) of abnormalities. As expected, common aneuploidies involving chromosomes 21, 18, 13, X, and Y were predominantly identified in: 82.7% (48/58) of fetuses and 41.9% (39/93) of POCs/IUDs/stillbirths (Figure 2a- c, Table 3, Figure 5). Rare autosomal aneuploidies accounted for 32.3% (30/93) and were observed exclusively in POCs. Similarly, triploidies (12.9%) were identified only in POCs, as expected.

**Figure 5:**
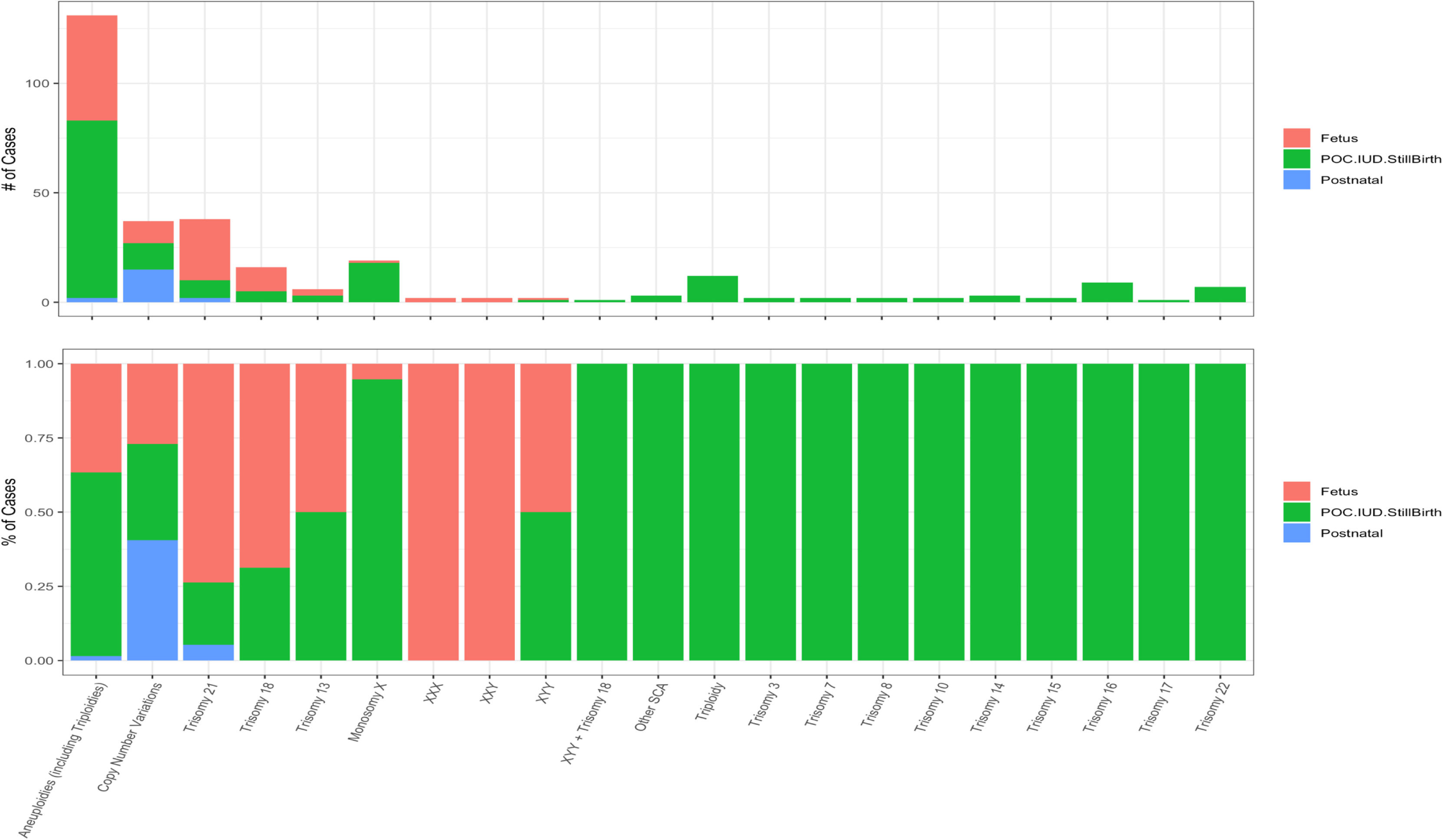
Aneuploidies and pathogenic and likely pathogenic variants identified by lpWGS

Pathogenic/Likely pathogenic CNVs - Smaller CNVs constituted 22% (37/168) of all cases. Smaller CNVs were detected in 12.9% (12/93) of POCs/IUDs/stillbirths (Figure 5). In postnatal samples, pathogenic CNVs were more common (88.2%, 15/17), while common aneuploidies were observed in only 12% of cases (Figure 5).. The pathogenic CNVs ranged in size from 1 Mb to 81.4 Mb. Of these: 8.3% (3/36) involved two variants per sample, while the remaining cases each showed a single variant. A total of 11.9% (20/168) of variants were <5 Mb, sizes that would have been missed by conventional karyotyping. One sample exhibited terminal gains and losses, suggesting a balanced translocation in the parents; parental testing was recommended, however this case was lost to follow-up.

Identification of common microdeletion syndromes – Multiple well known microdeletion syndromes were readily identified, including: 22q11.2 deletion syndrome (T11, T32, T35), Angelman/Prader-Willi syndrome (T24, T27), Williams syndrome (T8,T13,T31,T36), Cri-du- chat syndrome (T30), Jacobsen syndrome (T22,T33) These findings are detailed in (Figure 2d-g, Table 4).

**Table 4:** Summary of the smaller pathogenic copy number variation identified by the low pass whole genome sequencing. – First Trimester, 2^nd^ Trimester – Second Trimester.

| SI No. | Sample Type | Age Range | Cytoband | Size (Mb) | Associated Syndrome |
| --- | --- | --- | --- | --- | --- |
| T1 | Amniotic Fluid | 1<sup>st</sup> Trimester | Del:17p12 | 1.50 | Charcot-Marie-Tooth Disease Type 1A (CMT1A) |
| T2 | Amniotic Fluid | - | Del:17q11.2 | 2.60 | - |
| T3 | Amniotic Fluid | 2<sup>nd</sup> Trimester | Del:7q31.33-q33 | 10.50 | - |
| T4 | Amniotic Fluid | 2<sup>nd</sup> Trimester | Del:17q12 | 1.75 | Renal Cysts and Diabetes Syndrome (RCAD) |
| T5 | Chorionic Villus Sample | 1<sup>st</sup> Trimester | Mos.Del: Xq24 | 2.45 | - |
| T6 | Chorionic Villus Sample | - | Del:16p12.2-p11.2 | 8.00 | Rubinstein-Taybi Syndrome |
| T7 | Amniotic Fluid | 2<sup>nd</sup> Trimester | Del: Xp22.31 | 1.85 | Ichthyosis, X-linked (Steroid Sulfatase Deficiency) |
| T8 | Amniotic Fluid | - | Mos.Del: 7q11.22-q11.23 | 3.75 | Williams Syndrome |
| T9 | Chorionic Villus Sample | 2<sup>nd</sup> Trimester | Mos.Del: 13q32.3–q33.1 | 1.50 | - |
| T10 | Amniotic Fluid | 2<sup>nd</sup> Trimester | Del: 12q21.1-q21.33 | 15.60 | Noonan Syndrome (PTPN11 gene) |
| T11 | Product of conception | - | Del: 22q11.21 | 1.41 | DiGeorge Syndrome/Velocardiofacial Syndrome |
| T12 | Product of conception | - | Del: 15q26.2-q26.3 | 7.80 | - |
| T13 | Product of conception | - | Del: 7q11.21-q11.23 | 13.96 | Williams Syndrome |
| T14 | Product of conception | - | Mos. Dup: 5q23.1-q23.3 | 14.00 | Crohn's Disease (NOD2 gene) |
| T15 | Product of conception | - | Del:8p23.3-p11.1, Mos.Dup:8q12.3 - q24.3 | 45.14,81.4 | Langer-Giedion Syndrome; Burkitt Lymphoma |
| T16 | Product of conception | 1<sup>st</sup> Trimester | Del: 2q36.1-q36.3, Del: 11q22.3 - q23.1 | 6.2,1 | Potocki-Shaffer Syndrome |
| T17 | Product of conception | - | Del: 6p24.3-p23, Del: 7q31.32 - q31.33 | 5.8,1.3 | Cornelia de Lange Syndrome |
| T18 | Product of conception | - | Del: 2q37.2-q37.3 | 6.95 | - |
| T19 | Product of conception | - | Del: 14q12 | 1.60 | - |
| T20 | Product of conception | - | Del:1q21.1-q21.2 | 1.80 | TAR Syndrome |
| T21 | Product of conception | - | Dup: 17p12 | 1.40 | - |
| T22 | Peripheral Blood | 0-5Y | Del: 11q23.3-q25 | 14.06 | Jacobsen Syndrome |
| T23 | Peripheral Blood | 0-5 Y | Del: 11q23.3-q25 | 14.06 | Jacobsen Syndrome |
| T24 | Peripheral Blood | 0-5 Y | Del: 15q11.2-q13.1 | 5.00 | Prader-Willi Syndrome, Angelman Syndrome |
| T25 | Peripheral Blood | 26-30 Y | Del: 10q21.1 | 1.90 | - |
| T26 | Peripheral Blood | 0-5 Y | Del: 1p32.1-p31.3 | 3.10 | - |
| T27 | Peripheral Blood | 16-20 Y | Dup: 15q11.2-q13.1 | 7.29 | Prader-Willi Syndrome, Angelman Syndrome |
| T28 | Peripheral Blood | 0-5 Y | Del: 10q11.22-q11.23 | 5.90 | - |
| T29 | Peripheral Blood | 0-5 Y | Dup: 12p13.33-p13.31, Dup: 6q15 | 9.66,1.3 | - |
| T30 | Peripheral Blood | 6-10 Y | Del: 5p15.33-p14.1 | 25.41 | Cri-du-Chat Syndrome |
| T31 | Peripheral Blood | 6-10 Y | Del: 7q11.23 | 2.65 | Williams Syndrome |
| T32 | Peripheral Blood | 6-10 Y | Del: 22q11.21 | 1.30 | DiGeorge Syndrome, Velocardiofacial Syndrome |
| T33 | Peripheral Blood | 0-5 Y | Dup: 7p22.3-p21.3 | 8.36 | Greig Cephalopolysyndactyly Syndrome |
| T34 | Peripheral Blood | 26-30 Y | Dup: 9p24.3-p13.1 | 38.71 | Alagille Syndrome (JAG1 gene) |
| T35 | Peripheral Blood | 0-5 Y | Del: 22q11.21 | 2.20 | DiGeorge Syndrome, Velocardiofacial Syndrome |
| T36 | Peripheral Blood | 0-5 Y | Del: 7q11.23 | 1.60 | Williams syndrome |
| T37 | Product of conception | - | Dup:13q31.3-q34, Del: 18q21.33 - q23 | 24.7;17.96 | - |

Variant of uncertain significance (VOUS) - The size of the 36 VOUS ranged from 1.05 Mb to

9.9 Mb (Supplementary Table S2). Parental testing was conducted in 15 parents (5 couples), identifying the variant in 2 parents, resulting in an inheritance rate of 40% (2/5 couples). This additional information facilitated further refinement of variant classification as well as genetic counselling for the family.

## Discussion

We present the first large study on Indian clinical samples demonstrating the clinical-grade diagnostic utility of low-pass whole genome sequencing (lpWGS) for identification of aneuploidies and copy number variations (CNVs) in prenatal and postnatal samples. Our study achieved 100% detection rate of aneuploidies and CNVs previously detected by existing gold-standard technique such as chromosomal microarray (CMA). Additionally, we were able to identify CNVs below the resolution threshold of karyotyping, highlighting its superior resolution. Furthermore, we also implemented a method which led to successful identification of the chromosomal origin of imbalances in abnormal karyotypes, particularly in cases involving derivative chromosomes.

The rigorous validation process was divided into two-phases. The first phase was designed for detecting CNVs ≥1 Mb. Encouraged after obtaining 100% concordance we increased the resolution of LOD for CNV to 50 Kb in the second phase. This approach aligns with previous studies that reported reliable CNV detection at resolutions of 50–100 kb.^12,13^. Importantly, our data showed no difference in resolution for CNV detection between prenatal and postnatal samples, underscoring the robustness of lpWGS across diverse sample types.

The depth of sequencing (0.5–0.7X) employed in our study was optimized for cost- effectiveness and scalability while maintaining high diagnostic accuracy. At this shallow sequencing depth, lpWGS effectively detected whole chromosome aneuploidies and small to large structural abnormalities. Similar studies have demonstrated the reliability of lpWGS at depths as low as 0.25X (Dong et al., 2017)^11^, although depths below 0.5X may compromise the identification of triploidy and absence of heterozygosity.^10,19^ By increasing the sequencing depth to 3-4X in phase two, we successfully identified triploidies using a SNP-based algorithm, further enhancing the clinical utility of lpWGS.

Comparisons between CNV data derived from microarrays and lpWGS showed high concordance in the estimated sizes of chromosome imbalances, with minor discrepancies in terms of chromosomal location attributable to differences in probe spacing across platforms. At a resolution of 1 Mb, our overall diagnostic yield for pathogenic/likely pathogenic variants was 13.6%, comparable to the yield reported by Wang et al.^13^ By including VOUS, the yield increased to 16.3%, consistent with yields reported in prior studies.^10,11,12^ Diagnostic yields vary widely (2.83–53%) depending on sequencing depth, resolution, and sample types, with the highest yields observed in POCs.

In line with expectations, aneuploidies were the predominant abnormalities detected in fetuses (82.7%) and POC/stillbirth samples (41.9%), while smaller CNVs were more prevalent in postnatal samples (88.2%). The technique reliably identified well-known microdeletion syndromes, such as 22q11.2 deletion, Cri-du-chat syndrome, Angelman/Prader-Willi syndrome, 1p36 deletion syndrome, and Wolf-Hirschhorn syndrome.

We observed a VOUS rate of 2.9%, primarily in postnatal samples. It has been previously suggested that lpWGS tends to detect more VOUS compared to CMA due to improved resolution and uniform genome-wide coverage.^13^ Previous studies have reported VOUS rates ranging from 4–5.8% even by CMA. Therefore the rate observed in our study of 1260 samples by lpWGS was within the range reported in literature.^20^ As with other genetic tests, VOUS detection poses challenges for genetic counseling, particularly in prenatal settings. Guidelines recommend careful interpretation with parental segregation testing, cautious phenotypic correlation, and avoidance of reproductive decisions based solely on VOUS findings.^21^ Conversly, lpWGS may also identify novel clinically relevant CNVs that are undetectable by CMA.

Detection of mosaicism presents another challenge in clinical diagnostics. Mosaic aneuploidies are kown to occur in <1% of prenatal cases.^22^ Our data showed reliable mosaicism detection at levels 15%, however size variability was seen in one sample. That sample probably had a complex karyotype involving chromosome X. Previous studies have demonstrated mosaicism detection down to 5%.^9^ Mosaicism detection accuracy remains influenced by platform variability, sample quality, and maternal cell contamination. Clinical interpretation of mosaicism, particularly at low levels (15–20%), requires caution, as it may lead to diagnostic uncertainty and challenges in genetic counselling.^23^

Apart from the accuracy, our study also demonstrated that lpWGS has an extremely low assay failure rate of 0.2%, significantly lower than that reported for the CMA (∼4.6%).^10^ This low failure rate is particularly advantageous in prenatal settings, where re-sampling is often impractical. Furthermore, lpWGS requires low input DNA (25–50 ng) compared to CMA (300 ng), making it suitable for resource-limited scenarios. Cost-effectiveness is another major advantage of lpWGS assay. With lpWGS the cost of testing is reduced by 50% as compared to CMA, our own calculations showed a 40-50% reduction in cost compared to low resolution microarray.^10^ As sequencing costs continue to decline, lpWGS is expected to become increasingly accessible, particularly in low- and middle-income countries.^24^

One of the limitations of this study was that we did not evaluate additional rare variants seen on lpWGS. Literature suggests that this technique may identify novel variants undetectatble by microarrays and detected by lpWGS. Additionally, in phase two, we restricted our scope to triploidies and did not analyze regions of homozygosity/uniparental disomy, these are planned for subsequent phases at higher sequencing depth and resolution.

## Conclusion

In summary, lpWGS is a robust, sensitive, and scalable method for genome-wide detection of aneuploidies and pathogenic CNVs. It’s superior resolution, cost-effectiveness, and ability to detect a wide range of abnormalities makes it an excellent alternative to CMA, particularly in resource-limited clinical settings.

## Supporting information

https://docs.google.com/document/d/1VDDskknuaR1hi0w8Up9nfO0AqGMHIYU_7xFzK_qnjzI/edit?usp=sharing

## Data Availability

All data produced in the present study are available upon reasonable request to the authors

