## Supplementary material for "Low Pass Whole Genome sequencing - a cost-effective new technique for the identification of aneuploidies and copy number variations: a study of 1372 clinical samples in an Indian cohort": https://docs.google.com/document/d/1VDDskknuaR1hi0w8Up9nfO0AqGMHIYU_7xFzK_qnjzI/edit?usp=sharing

**Supplementary** **Tables**

**Table S1: Reproducibility analysis performed on 13 samples**

| **Sl No** | **Sample Type** | **Cytoband** | **Run I** | | | **Run II** | | | **Run III** | | |
| --- | --- | --- | --- | --- | --- | --- | --- | --- | --- | --- | --- |
|  |  |  | **Start** | **End** | **Size (Mb)** | **Start** | **End** | **Size (Mb)** | **Start** | **End** | **Size (Mb)** |
| V35 | AF | Del:15q13.2q13.3 | 30786834 | 32089069 | 1.3 | 30786834 | 31976316 | 1.2 | 30885000 | 32485000 | 1.6 |
| V14 | AF | Del:3q29 | 193046654 | 195610869 | 2.6 | 192919987 | 195353649 | 2.4 | 192801721 | 195307011 | 2.5 |
| V29 | AF | Del:9q22.1q22.31 | 87944362 | 92695235 | 4.8 | 88064703 | 92809363 | 4.7 | 90578812 | 95478796 | 4.9 |
| V30 | AF | Del:10p15.3p14 | 1 | 7581645 | 7.6 | 1 | 7581645 | 7.6 | 0 | 7,760,000 | 7.7 |
| V25 | AF | Del:6q26q27; Dup:7q33q36.3 | 161600873; 133695914 | 170394097; 159307878 | 8.8; 25.6 | - |  |  | 162635406;  133213445 | 171115067;  159138663 |  |
| V23 | AF | Del:5p15.33p15.2;  Dup: 20p13p12.1 | 1; 1 | 13808660; 15504001 | 13.8; 15.5 | 1 | 13808660 | 13.8 | 1410000; 0 | 13810000;  15460000 | 12.4;  15.5 |
| V46 | PB | Del:22q13.32q13.33 | 48152115 | 50561615 | 2.4 | 48152115 | 50561615 | 2.4 | 48470380 | 51304566 | 2.8 |
| V27 | AF © | Del:7p21.3p15.2 | 10612959 | 26458168 | 15.8 | 10612959 | 26458168 | 15.8 | 10660000 | 26560000 | 15.9 |
| V2 | CVS | Trisomy 18 | 1 | 80140651 | 80.1 | 1 | 80140651 | 80.1 | 0 | 78077248 | 78.1 |
| V1 | CVS | Trisomy 21 | 13850829 | 46484554 | 32.6 | 13850829 | 46484554 | 32.6 | 0 | 48129895 | 48.1 |
| V9 | POC | Trisomy 12 | 1 | 133110963 | 133.1 | 1 | 133110963 | 133.1 | 0 | 133851895 | 133.9 |
| V32 | POC | Dup:11p15.5q23.3; Del:22q11.1q11.21 | 3619629; 16635150 | 116866114; 20555596 | 113.2; 3.9 | 423669; 16635150 | 116866114; 20268189 | 116.4; 3.6 | 0;  17062659 | 135006516;  20970380 | 135.0;  3.9 |
| V34 | POC | Del:13q32.3q34; Dup:16p13.3p13.2 | 101143327; 5061519 | 113398857; 8815533 | 12.3;  3.8 | 101143327; 1 | 114122514; 8815533 | 13.0 8.8 | 101710324;  31910000 | 115169878;  34489689 | 13.5;  2.6 |

**Table S2: Variant of Unknown Significance (VoUS) Cases**

| **Sl. No** | **Sample Type** | **Gestation Age /Age** | **Cytoband** | **Size (Mb)** |
| --- | --- | --- | --- | --- |
| ST1 | Amniotic Fluid | First Trimester | Dup: 18p11.32-p11.31 | 1.70 |
| ST2 | Amniotic Fluid | - | Dup: 9p21.3 | 2.00 |
| ST3 | Amniotic Fluid | - | Dup: Xp22.31 | 2.15 |
| ST4 | Amniotic Fluid | - | Dup: 19p12 | 1.00 |
| ST5 | Amniotic Fluid | - | Dup: 9p23 | 1.20 |
| ST6 | Amniotic Fluid | - | Dup: 6p12.1-q11.1 | 5.53 |
| ST7 | Amniotic Fluid | - | Dup: 20p12.2-p12.1 | 7.40 |
| ST8 | Amniotic Fluid | - | Del: 5q13.1-q13.2 | 2.40 |
| ST9 | Amniotic Fluid | Second Trimester | Dup: 1p31.1 | 2.20 |
| ST10 | Amniotic Fluid | Second Trimester | Dup: 8p12 | 1.00 |
| ST11 | Amniotic Fluid | Second Trimester | Del: 5q12.1 | 2.00 |
| ST12 | Amniotic Fluid | Second Trimester | Dup: 6p23-p22.3 | 1.60 |
| ST13 | Amniotic Fluid | First Trimester | Dup: 16q23.3 | 1.70 |
| ST14 | Amniotic Fluid | Second Trimester | Dup: 5p15.31-p15.2 | 1.60 |
| ST15 | Amniotic Fluid | Second Trimester | Del: 1p36.22-p36.21 | 1.70 |
| ST16 | Amniotic Fluid | - | Dup: 2q21.1 | 1.00 |
| ST17 | Amniotic Fluid | Second Trimester | Dup: 15q13.2-q13.3 | 2.70 |
| ST18 | Amniotic Fluid | Second Trimester | Dup: 4q28.1 | 1.50 |
| ST19 | Amniotic Fluid | Second Trimester | Del: 3p26.3-p26.2 | 2.20 |
| ST20 | Amniotic Fluid | Second Trimester | Dup: 7q35-q36.1 | 1.60 |
| ST21 | Chorionic Villus Biopsy | - | Dup: 2q31.3-q32.1 | 2.20 |
| ST22 | Product of Conception | - | Del: 8p23.1 | 1.05 |
| ST23 | Product of Conception | - | Del: 11p15.4-p15.2 | 2.40 |
| ST24 | Product of Conception | - | Del: 5q13.1-q13.2 | 2.60 |
| ST25 | Product of Conception | - | Dup: 14q31.1 | 1.00 |
| ST26 | Product of Conception | - | Del: 15q11.2 | 2.94 |
| ST27 | Product of Conception | - | Dup: 15q13.2-q13.3 | 2.70 |
| ST28 | Product of Conception | - | Dup: 22q11.23 | 1.50 |
| ST29 | Product of Conception | - | Del: 1p36.22-p36.21 | 1.40 |
| ST30 | Peripheral Blood | 0-5Y | Dup: Xp22.31 | 1.85 |
| ST31 | Peripheral Blood | 31-35Y | Dup: Xp22.31 | 1.75 |
| ST32 | Peripheral Blood | 0-5Y | Dup: Xp22.31 | 1.75 |
| ST33 | Peripheral Blood | 36-40Y | Dup: 6p23-p22.3 | 1.40 |
| ST34 | Peripheral Blood | 0-5Y | Dup: 9p23-p21.3 | 9.90 |
| ST35 | Peripheral Blood | 0-5Y | Dup: 13q31.2 | 1.40 |
| ST36 | Peripheral Blood | 26-30Y | Dup: 7q35q36.1 | 1.60 |
